# COVID-19 Vaccine Prioritisation in Japan and South Korea

**DOI:** 10.1101/2021.04.16.21255649

**Authors:** June Young Chun, Hwichang Jeong, Philippe Beutels, Norio Ohmagari, Yongdai Kim, Shinya Tsuzuki

**Affiliations:** Department of Internal Medicine, National Cancer Center, Goyang, South Korea; Department of Statistics, Seoul National University, Seoul, South Korea; Faculty of Medicine and Health Sciences, University of Antwerp, Antwerp, Belgium; Disease Control and Prevention Center, National Center for Global Health and Medicine, Tokyo, Japan; AMR Clinical Reference Center, National Center for Global Health and Medicine, Tokyo, Japan

## Abstract

**Background:** Due to a limited initial supply of COVID-19 vaccines, the prioritisation of individuals for vaccination is of utmost importance for public health. Here, we provide the optimal allocation strategy for COVID-19 vaccines according to age in Japan and South Korea.

**Methods:** Combining national case reports, age-specific contact matrices, and observed periods between each stages of infection (Susceptible-Exposed-Infectious-Quarantined), we constructed a compartmental model. We estimated the age-stratified probability of transmission given contact (*q*_*i*_) using Bayesian inference method and simulated different vaccination scenarios to reduce either case numbers or death toll. We also performed sensitivity analyses on the proportion of asymptomatic cases and vaccine efficacy.

**Findings:** The model inferred age-stratified probability of transmission given contact (*q*_*i*_) showed similar age-dependent increase in Japan and South Korea. Assuming the reported COVID-19 vaccine efficacy, our results indicate that Japan and South Korea need to prioritise individuals aged 20–35 years and individuals aged over 60 years, respectively, to minimise case numbers. To minimise the death toll, both countries need to prioritise individuals aged over 75 years. These trends were not changed by proportions of asymptomatic cases and varying vaccine efficacy on individuals under 20 years.

**Interpretation:** We presented the optimal vaccination strategy for Japan and South Korea. Comparing the results of these countries demonstrates that not only the effective contact rates containing *q*_*i*_ but also the age-demographics of current epidemic in Japan (dominance in 20s) and South Korea (dominant cases over 50s) affect vaccine allocation strategy.

## Introduction

The pandemic of coronavirus disease 2019 (COVID-19), an infectious disease caused by severe acute respiratory syndrome coronavirus 2 (SARS-CoV-2), is still ongoing in a rapid and a widespread manner. The high transmissibility of SARS-CoV-2 might be attributed to its route of transmission (respiratory) and its viral shedding pattern (peak viral loads at the symptom onset).^1,2^ These characteristics hinder efficient public health control of COVID-19.

Therefore, COVID-19 vaccines are expected to be a game changer for ending this pandemic. Several promising vaccines are under development or have received emergency use authorisation in many countries including the United Kingdom (UK) and the United States (US).^3-5^

Vaccine supply, however, is expected to be limited in 2021. As of March 31, 2021, Japan immunized 877,159 individuals (0·7%) and South Korea immunized 852,202 individuals (1·6%) at least the first dose, which are far less proportions than countries in Europe and America.^6,7^ Determination of who to prioritise for COVID-19 vaccination is an important matter that plays a major role in curbing the growing burden of the disease.

Vaccinating the younger population, who have high contact rates, could lower the total COVID-19 incidence, whereas targeting the elderly, who have high mortality rates, would lower the death toll.^8,9^ Given the scarcity of vaccines, the World Health Organisation (WHO) recommended to focus initially on direct reduction of mortality, and maintenance of most critical essential workers.^10^ Likewise, many countries including the European Union, UK, and the US announced interim recommendations in which healthcare personnel, long-term care facility residents, and elderly people with various age cut-offs were prioritised for COVID-19 vaccination.^11,12^ By contrast, Indonesia plans to vaccinate working young adults before the elderly.^13^ Indeed, the vaccination strategy could vary in countries, and the optimal allocation strategy could be informed by modelling studies accounting for country-specific characteristics.

Here, we constructed the modified Susceptible-Exposed-Infectious-Quarantine (SEIQ) model, using the observed distributions of the latent period (E → I) and the diagnostic delay (I → Q), along with the transmission onset distribution relative to the symptom onset.^14^ Bayesian inference with Markov Chain Monte Carlo method was used for parameter estimation of age-stratified force of infection (S → E). By virtue of this realistic compartmental model, the main objective of this study is to determine the optimal administration of COVID-19 vaccines, in both Japan and South Korea, in terms of reduction of (a) total incidence and (b) the death toll.

## Methods

### Epidemiologic data

The epidemiologic data consisted of COVID-19 reports provided by the Ministry of Health, Labour and Welfare of Japan and the Ministry of Health and Welfare of South Korea.^6,7^ Both governments report COVID-19 cases by 10-year age interval, diagnosed with SARS-CoV-2 polymerase chain reaction test. We could regard those numbers as real case numbers according to the ascertainment levels close to 100% in Japan and South Korea, given the baseline case-fatality ratio of 1·4%.^15^ The age-specific mortality ratios were also collected.^6,7^

### Demography and contact matrix

Age-structured population data were obtained from the Statistics Bureau Japan and Statistics Korea.^16,17^ We introduced contact matrices in our compartmental model, using the contact survey data by Ibuka et al. for Japan, and projected contact matrix by Prem et al. for South Korea.^8,18^ New contact matrix with 5-year age intervals was established for Japan from diary data that included 4331 respondents using a bootstrap method.^18,19^

To capture the changes of contact patterns as a result of social distancing measures, we took into account school closure policies and reduced contact rates at both work and other places using Google mobility data.^20-22^ In Japan, most schools that had been temporarily closed reopened on June 1, 2020, whereas in South Korea, school attendance has been capped at two-thirds during national distancing level 1, one-thirds during level 2 (except for high schools which remained 2/3), and remote learning only during level 3 throughout 2020 (Figure 1).

**Figure 1.**
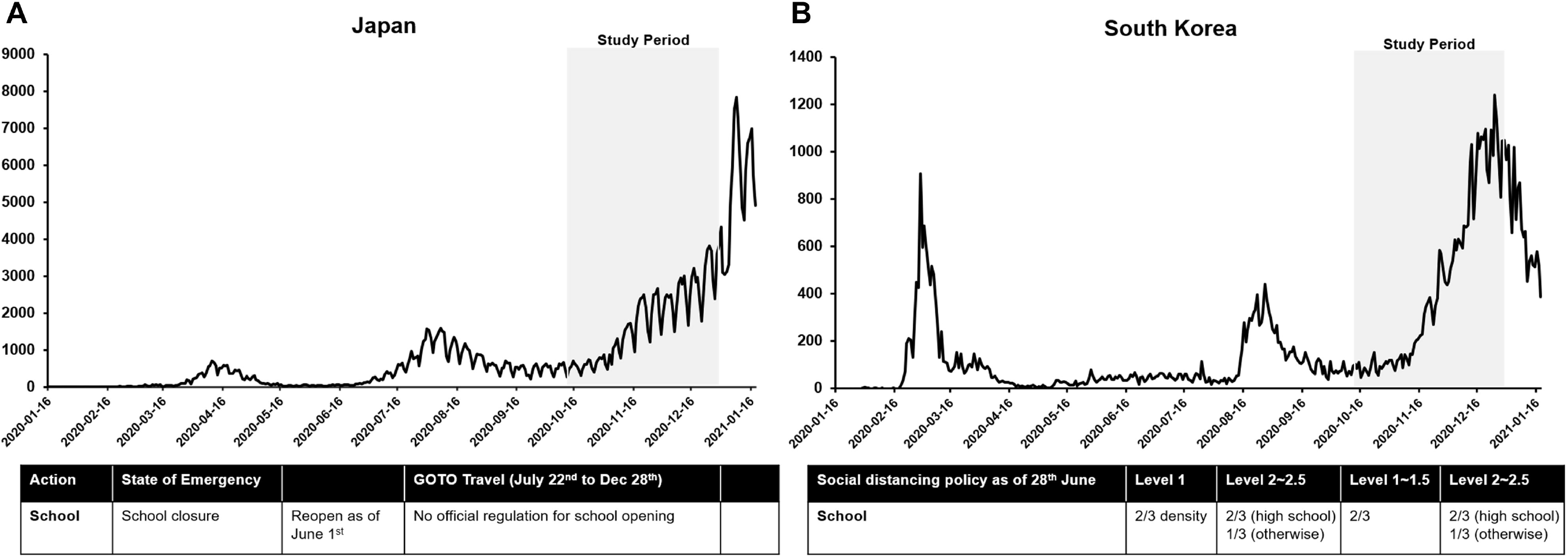
Epidemic curve and national intervention in (A) Japan and (B) South Korea.

### Transmission model

What we could observe is the age-stratified (10-year interval) incidence of COVID-19. This was made compatible to the contact matrix by dividing the incidence data in 5-year instead of 10-year age groups proportionally to demographic structure. From previous epidemiologic studies, we know the (i) incubation period distribution, (ii) transmission onset distribution relative to symptom onset, and the (iii) delay from symptom onset to diagnosis.^14^ The (iv) latent period distribution could be derived from both incubation period and transmission onset distribution (Figure 2). In Japan and South Korea, individuals who have been diagnosed with COVID-19 are isolated immediately; thus, the confirmation date could be regarded as the date on which isolation had begun.

**Figure 2.**
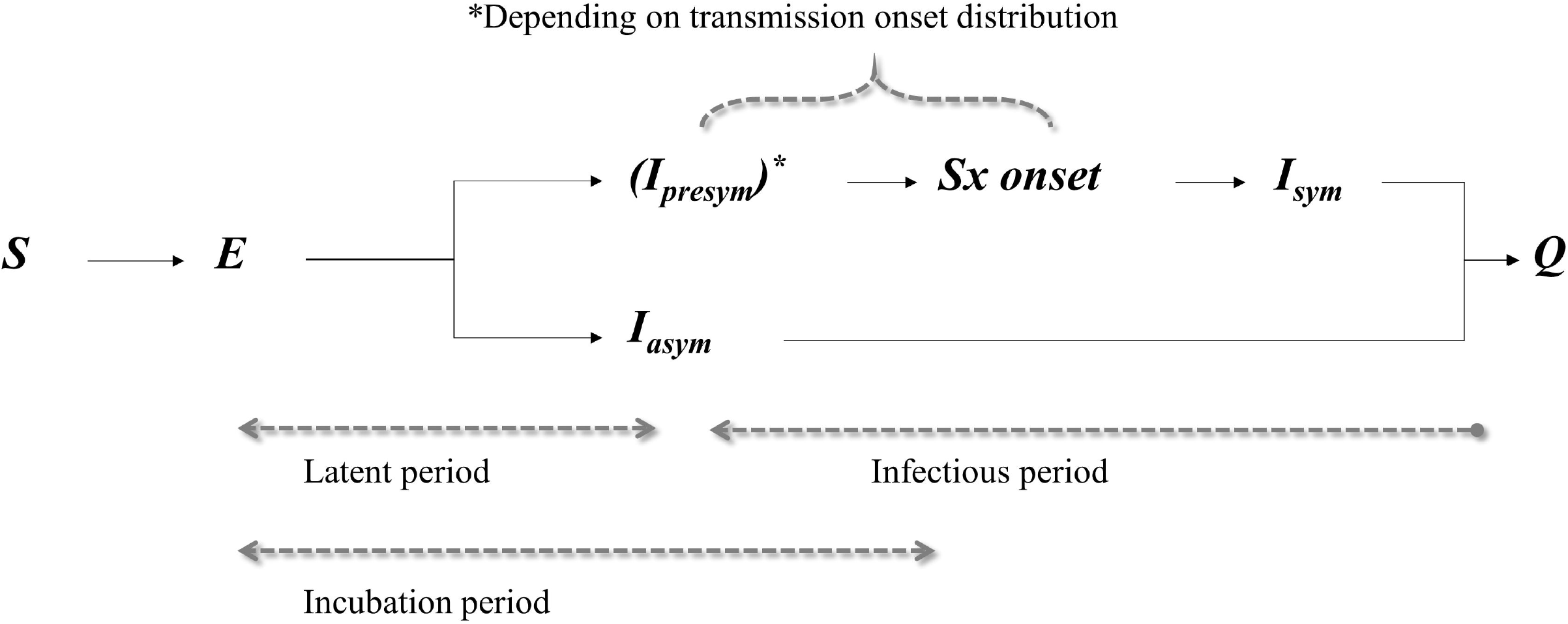
A schematic plot of different time periods of the transmission of SARS-CoV-2.

For those who eventually developed symptoms (*Ipresym\** → *Isym*), we could infer the individual’s symptom onset relative to the diagnosed date using (iii). Then, we could successively infer transmission onset using (ii) and exposure date using (i).

For those who never developed any symptoms (*Iasym*), accounting for 16% of total infection by a meta-analysis^23^, we assumed that their (iv) latent period distribution is the same as those who developed symptoms. Also, we regard that the infectious period distribution for *Iasym* is the same as the total infectiousness period of symptomatic cases as suggested.^24^ We suppose that the *Iasym* is quarantined at random during the infectious period, and the relative infectiousness of the *Iasym* is half of that of the others.^24^

Then, what is left to estimate is the age-stratified force of infection (S → E). According to Vynnycky and White,^25^ the force of infection *λ*_*i*_ experienced by age group *i* is:

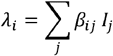

Here, *β*_*ij*_ is the rate at which susceptible in the age group *i* and infectious individuals in the age group *j* come into effective contact per unit time, and *I*_*j*_ is the number of infectious individuals in the age group *j*. We further divide *β*_*ij*_ into:

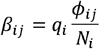

Here, *q*_*i*_ is the probability that a contact between a susceptible in age group *i* and an infectious person leads to infection, *ϕ*_*ij*_ is the number of contacts an individual of age group *j* makes with those of age group *i* per unit time, and *N*_*i*_ is the number of individuals in age group *i*. Since we know the contact matrix for Japan and South Korea, we could estimate *q*_*i*_ for both the countries. All analyses were conducted using the *R* statistical software version 3.6.3. Detailed Bayesian inference methods are available in the Supplementary Method. Code and data to reproduce the analyses are available as an *R* package at *https://github.com/Hwichang/COVID-19-Vaccine-Prioritisation*.

Japan and South Korea have similar epidemiological trends; (a) the import-driven first wave, (b) domestic origin second wave closely related to nightlife clusters,^26,27^ and (c) the third wave possibly enhanced by a seasonality factor (Figure 1). We used the third wave mainly for our simulation considering that this period could reflect the real characteristics of SARS-CoV-2 transmission in each country not only excluding the importation factor, but also including the effects of seasonality similar to other respiratory viruses. Thus, the study period was set from October 15^th^ to December 25^th^, 2020, for Japan prior to the winter vacation, and from October 15^th^ to December 22^nd^, 2020, for South Korea prior to the “ban of gathering of five or more people”. During the third wave, people in their 20s still constituted most of the confirmed cases in Japan following the second wave, whereas in South Korea, this shifted to those over 50 during the third wave.

### Vaccination scenarios

Using the SEIQ model, we could calculate how many cases and deaths would arise without any intervention, including vaccination, after 2 weeks of the study period by means of forward projection. We set this as the baseline scenario and explored how to allocate COVID-19 vaccines according to age groups, with stepwise increase in vaccine supply relative to the population size. Being vaccinated, the susceptible population in each age group would decrease, and our ultimate goal was to find the optimal distribution of vaccines to reduce COVID-19 cases and deaths.

### Sensitivity analysis

Among model parameters, proportion of asymptomatic cases and vaccine efficacy were hypothesised values. We thus tried to vary those values with sensitivity analysis. For the proportion of asymptomatic cases, the meta-analysis showed that it ranged from 4% to 41%, and we varied the parameter accordingly from 4% to 40%.^23^ As far as vaccine efficacy was concerned, we adopted the age-dependent COVID-19 vaccine efficacy with statistical significance.^4^ Since there is a paucity of information on vaccine efficacy of children, we first assumed the same efficacy as young adults, and further analysed the sensitivity with half efficacy of children. Lastly, we assumed the COVID-19 vaccine efficacy against asymptomatic infection as 70% across all analyses.^28^

### Ethics approval

All the data used in this study were publicly available, and therefore were regarded exempt from institutional review board assessment.

## Results

### Model estimation

The age-stratified probability (*q*_*i*_) that a susceptible person of age group *i* acquires infection given contact with an infectious person was estimated as in Figure 3. Both Japan and South Korea showed similar trends of age-dependent increase. The lowest *q*_*i*_ value was seen in the age group of 5 to 10 years (0·0043, 95% CI [0·0040–0·0045] in Japan and 0·0293, 95% CI [0·0270–0·0316] in South Korea), and the highest value was seen in the age group of over 75 years (0·1599, 95% CI [0·1568–0·1628] in Japan and 0·1577, 95% CI [0·1518– 0·1637] in South Korea). The model parameters are summarised in Table 1.

**Figure 3.**
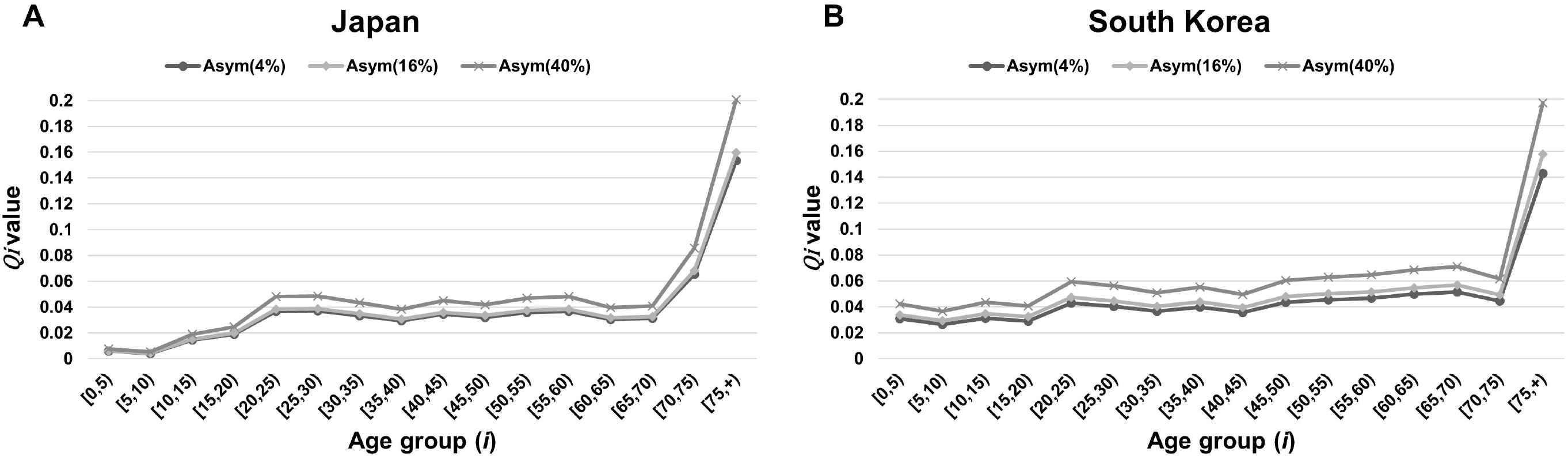
The age-stratified probability of transmission given contact (*q*_*i*_) of COVID-19 in (A) Japan and (B) South Korea. As a sensitivity analysis, the *q*_*i*_ values with varying proportions of asymptomatic cases from 4% to 40% were presented together.

**Table 1.**
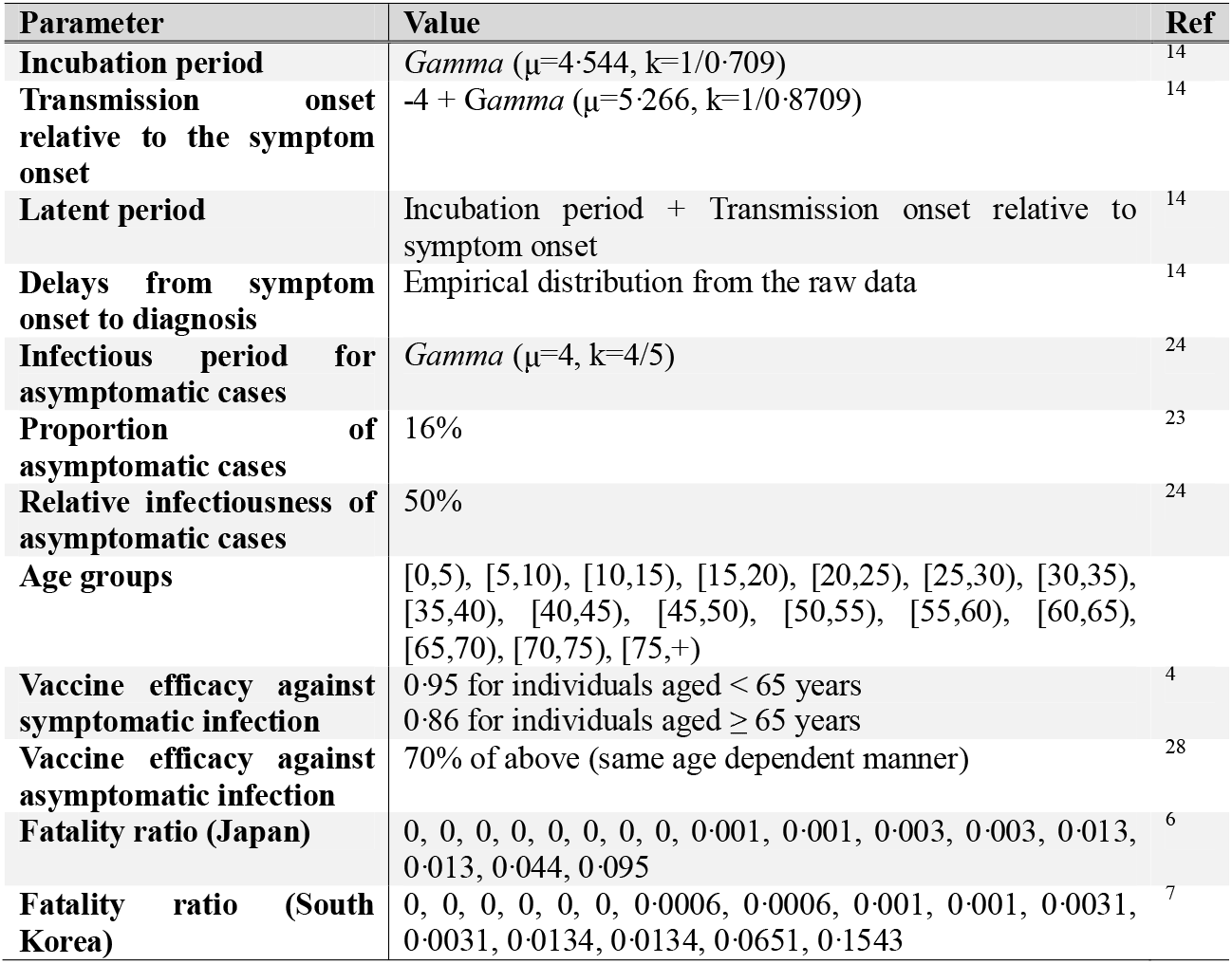
Model parameters.

### Vaccine allocation by age groups

With a provision of 20% to 100% of COVID-19 vaccines, considering the population size of Japan, the optimal vaccine allocation to reduce total case and death numbers are shown in Figure 4A. With a provision of 20% to 100% of COVID-19 vaccines, considering the population size of South Korea, the optimal vaccine allocation to reduce total case and death numbers are shown in Figure 4B. The very first group to receive vaccination to reduce case numbers is aged 20–35 years in Japan and aged over 60 years in South Korea. In the same manner, the first group to receive vaccination to reduce the death toll is aged over 75 years in both countries.

**Figure 4.**
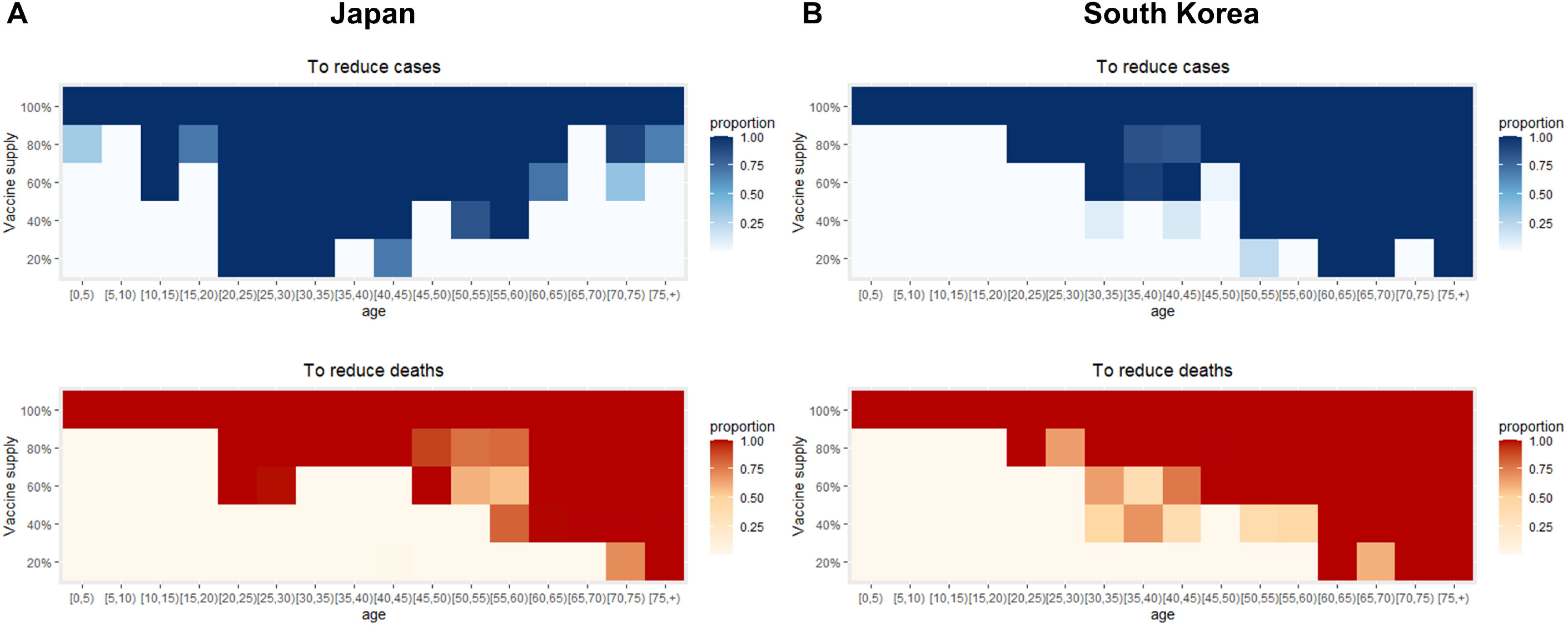
The optimal allocation strategy of COVID-19 vaccines according to age in (A) Japan and (B) South Korea, in terms of case and death reduction.

### Reduction in the number of cases and deaths

Assuming that there were no further intervention strategies including vaccination, during the third wave, there might have been 75,767 cases and 1,046 deaths in Japan and 25,521 cases and 529 deaths in South Korea in the following 2 weeks of the study period. With gradual vaccination in both countries, the lowest number of cases and deaths are shown in Table 2.

**Table 2.** The lowest expected number of COVID-19 cases and deaths at 2 weeks after the third wave by stepwise vaccine supply.

| Nation | Vaccine supply |  |  |  |  |  |
| --- | --- | --- | --- | --- | --- | --- |
| Japan | No Vaccine | 20% | 40% | 60% | 80% | 100% |
| Cases | 75 767 | 40 273 | 22 870 | 13 120 | 7266 | 3468 |
| (Range*) | (72 917–79 759) | (38 813–41 777) | (22 091–23 683) | (12 675–13 464) | (6956–7437) | (3404–3567) |
| Deaths | 1046 | 306 | 153 | 117 | 86 | 76 |
| (Range*) | (1007–1084) | (293–317) | (152–159) | (110–118) | (83–93) | (74–78) |
| South Korea | No Vaccine | 20% | 40% | 60% | 80% | 100% |
| Cases | 25 521 | 16 090 | 9923 | 5634 | 2558 | 1059 |
| (Range*) | (23 468–27 774) | (14 830–17 469) | (9169–10749) | (5218–6035) | (2416–2711) | (1002–1102) |
| Deaths | 529 | 94 | 67 | 47 | 38 | 33 |
| (Range*) | (488–572) | (88–103) | (62–68) | (45–49) | (35–39) | (31–34) |

\* Range was calculated based on the 95% Confidence interval of $q_i$ values.

### Sensitivity analysis

Varying the proportion of asymptomatic cases from 4% to 40%, vaccine priority groups were not changed in terms of case and death reduction. Likewise, reducing the vaccine efficacy of individuals under 20 years to 50% did not change the priority, either (Supplementary Figure 1).

## Discussion

Given the limited initial availability of COVID-19 vaccines, the prioritisation of individuals for vaccination is of utmost importance to public health workers and policy makers. Based on mathematical modelling, we hereby present estimates to inform the distribution of COVID-19 vaccines among different age groups, both in Japan and South Korea, with a stepwise increase in vaccine supply. Applying the reported COVID-19 vaccine efficacy,^4^ the initial target groups to reduce the total incidence were young adults aged 20–35 years in Japan and individuals aged over 60 years in South Korea. We interpret that this difference might be attributed to the different age-group distribution of COVID-19 cases between Japan (dominance in 20s) and South Korea (dominance over 50s) during the third wave, since the force of infection *λ*_*i*_ is not only affected by *β*_*ij*_, but also by *I*_*j*_ which is the number of infectious individuals in the age group. On the other hand, the target groups to reduce total deaths were primarily focused on the elderly, aged over 75 years in both countries.

To achieve the best model of SARS-CoV-2 transmission, we constructed a compartmental model based on a solid epidemiological investigation which could increase the reliability of our study.^6,7,14^ The estimated age-stratified probability of transmission given contact (*q*_*i*_) revealed similar results with other studies showing that children have lower probability of transmission given contact.^29^ However, it is yet to be elucidated whether children are less susceptible to COVID-19 or they show fewer clinical symptoms than adults, or both. To specify the relative impact of those factors, a well-designed national seroprevalence study would be necessary. In Japan and South Korea, seroprevalence of SARS-CoV-2 specific antibodies were measured in local regions including the capital city from May to June 2020, and the results ranged from 0·03% to 0·17% in Japan and 0·07% in South Korea. Owing to a low seroprevalence in both countries, it was not appropriate to derive an answer to the above question. Moreover, there is a caveat to interpret the *q*_*i*_ results, since it could be affected by contact numbers, *ϕ*_*ij*_. Japan showed much lower *q*_*i*_ values for individuals aged below 10 years than South Korea, and we assumed that the different school closure policies could explain the discrepancy; contrary to Japan, South Korea maintained restrictions on the number of students in school and the *q*_*i*_ might have decreased as low as Japan if it had not regulated school attendance. In addition, previous studies suggested that older age might contribute to vulnerability to COVID-19 infection, it is possible that biological aspects also affect the value of *q*_*i*_ then further investigation would be desirable in this area.^30^

There are several limitations to this study. First, although the contact matrix is the most important variable in this model, we do not have empirical data for the current mixing patterns. We tried to modify the existing prepandemic contact matrix to make it as realistic as possible, but it might be insufficient to reflect reality. Besides, in the existing Japanese contact matrix, the 20–29 year age group is marginally under-represented, and for South Korean contact matrix, we had to adopt the estimated result, not the measured one, since there was no available national survey data so far.^8,18^ Secondly, we did not take essential worker status and co-morbidities into consideration for our model. Third, seasonality would undoubtedly affect COVID-19 transmission, but it was hard to include this factor in our model. Then, our estimation might not be sufficient for prediction in future seasons. Fourth, the geographical heterogeneity was not considered in this study. In other words, differences in frequency of contact between urban and rural areas could not be assessed here. Lastly, the exact proportion of asymptomatic cases is still unelucidated. We adopted the results from a meta-analysis and further conducted a sensitivity analysis with variable ranges of asymptomatic proportion.^23^

Here, we present a realistic compartmental model for COVID-19 transmission based on the reliable epidemiological investigation conducted in two East Asian countries, Japan and South Korea. Furthermore, we provide the optimal distribution strategy of COVID-19 vaccines under limited vaccine supply in both countries. In short, the balance between age-dependent effective contact rates and age-specific incidences should be considered for national vaccination strategy. We believe that this study could guide public health practitioners based on the solid mathematical model.

## Supporting information

Supplementary description of Methods

## Data Availability

All the data and R codes used in this study are publicly available.

https://github.com/Hwichang/COVID-19-Vaccine-Prioritisation

## Author Contributions

JYC, YK and ST conceived and design the study. JYC and ST collected study data. HJ and YK analysed the data and JYC, PB, NO, and ST interpreted the results. JYC and HJ wrote a draft of the manuscript. All authors critically reviewed and approved the final manuscript.

## Acknowledgements

Japanese contact matrix data was kindly provided by Dr. Yoko Ibuka, Faculty of Economics, Keio University. We would like to thank Editage (www.editage.co.kr) for English language editing.

## Figure legends

**Supplementary figure 1. The optimal allocation strategy of COVID-19 vaccines according to age in Japan and South Korea with varying proportions of asymptomatic cases and vaccine efficacy**.

## Research in context Evidence before this study

We searched PubMed and MedRxiv for research articles published in English from database inception until March 11, 2021, with the keywords “SARS-CoV-2”, “COVID-19”, “Vaccine”, “Immunization,” “Distribution”, “Allocation”, and “Mathematical model”. We found several articles analysing the optimal distribution of COVID-19 vaccines fitted to each nation’s characteristics, notably in the US, UK, Germany, India, and South Korea. However, we identified no research identifying age-specific force of infection to COVID-19 in East Asia, further suggesting the optimal vaccination policy upon the mathematical model.

## Added value of this study

Our study combined solid epidemiological data from Japan and South Korea and built the reliable compartment model for SARS-CoV-2 transmission. Using the age-specific incidence of COVID-19, we could estimate the age-stratified probability of transmission given contact by Bayesian inference method, which was incorporated into the compartment model. By virtue of this model, we could evaluate the effects of vaccine distribution scenarios by age in terms of case and death reduction.

## Implications of all the available evidence

Fortunately, safe and effective COVID-19 vaccines have been developed within an unprecedented period. For now, how to implement vaccination most effectively is of utmost importance. Here, we suggest the vaccine prioritisation strategy in Japan and South Korea, and we believe it could be generalised to those who have a similar socio-demographic background.

