## Supplementary description of Methods for "COVID-19 Vaccine Prioritisation in Japan and South Korea"

### Supplementary method of COVID-19 Vaccine Prioritisation in Japan and South Korea

#### 1. BAYESIAN INFERENCE

##### A. overview

In a Susceptible-Exposed-Infectious-Recovered (SEIR) model, the force of infection $\lambda$, the rate at which susceptible individuals are infected (i.e exposed), is a crucial factor. The age-specific force of infection $\lambda_i$ in age group $i$ at discrete time $t$ could be written as:

$$\lambda_i = p(t|q_i) = \frac{q_i \sum_{j=1}^{A} \phi_{ij}(t)(I_{sym}^{j}(t-1) + 0.5 I_{asym}^{j}(t-1))}{n_i} \tag{S1}$$

where $q_i$ is the probability that a contact between a susceptible in age group $i$ and infectious person leads to infection, $\phi(t) = (\phi_{ij}(t))$ means contact matrix at discrete time $t$ ($\phi_{ij}(t)$ is the number of contacts an individual of age group $j$ makes with those of age group $i$ per unit time at discrete time $t$), $n_i$ is the number of individuals in age group $i$. $I_{sym}^{j}(t)$ and $I_{asym}^{j}(t)$ are the number of infectious individuals who either have symptom or not at discrete time $t$. And we suppose that the relative infectiousness of the $I_{asym}$ is half of $I_{sym}$. Here $i$ is in age group $\mathcal{A} = \{1, 2, \cdots, A\}$.

We try to estimate $q_i$ during the third wave in Japan and South Korea and denote it as $\theta_i$ hereafter. If we could observe the number of exposed individuals at time $t$ for age group $i$ and the number of total infectious individuals, the likelihood of the parameters could be easily derived. However what we could observe is only the number of diagnosed(i.e quarantined) individuals for age group $i \in \mathcal{A}$ at time $t$. In addition, asymptomatic infection which is the notable feature of COVID-19 should also be reflected in model. To resolve these difficulties, we use a Bayesian approach. In particular, we develop an efficient MCMC (Markov Chain Monte Carlo) algorithm in which the exposed date, symptom onset date and transmission onset date for all quarantined individuals are imputed with an assumption that there are 16% asymptomatic individuals. We explain the details of our Bayesian method in the following three subsections.

##### B. Data, Model and Posterior

The data we used in the analysis is daily numbers of quarantined individuals for each age group from January 2020 to January 2021. To estimate $\theta_i$, we are going to impute the exposed dates, symptom onset dates and transmission onset dates of all quarantined individuals conditional on given quarantined dates. For this purpose, we need a probability model which relates the exposed dates, symptom onset dates and transmission onset dates to the quarantined dates.

| Symptomatic cases | | Asymptomatic cases | |
|---|---|---|---|
| Symbol | Definition | Symbol | Definition |
| $E$ | Exposed date of an individual | $E$ | Exposed date of an individual |
| $Y$ | Incubation period | $L$ | Latent period |
| $I$ | Transmission onset time relative to the symptom onset | $C$ | Infectious period suspended by quarantine |
| $D$ | Diagnostic delay (i.e Quarantined) from the symptom onset | $R$ | Infectious period |

For each symptomatic individual, the quarantined date is sum of the exposed date($E$), the incubation period($Y$) and the period for symptom onset to diagnostic delay($D$). In addition, these individuals start infecting other susceptibles from the transmission onset date $(E + Y + I)$.

For each asymptomatic individual, the quarantined date is sum of exposed date ($E$), latent period($L$) and period for transmission onset to quarantined date($C$). We assume that the latent period distribution of asymptomatic individuals is same as that of symptomatic individuals.

Also the quarantined time distribution of $C(f_C)$ of asymptomatic individuals is set to be the exponential distribution with mean 1/1.7, which satisfies $P(C > R) = 0.01$, where $E + L + R$ is defined as the recovered date.

Finally, the quarantined date $T$ is defined as

$$T = (E + Y + D)\mathbb{1}(\Delta = 0) + (E + L + C)\mathbb{1}(\Delta = 1), \tag{S2}$$

where $\Delta$ is equal to 0 when the individual is symptomatic and 1 when asymptomatic. We set $\Pr(\Delta = 1) = 0.16$.

For individual $k$, let $W_k = (E_k, Y_k, I_k, L_k, C_k, D_k)$ and $V_k = (Y_k, I_k, L_k, C_k, D_k)$. Let $\mathcal{D}$ be the observed data which consist of the daily numbers of quarantined individuals. Our strategy to estimate $\theta_i$ is to generate $W_k$ and $\theta$ iteratively from their conditional posterior distributions $P(\theta_i|W, \mathcal{D})$ and $P(W_k|W_{(-k)}, \theta, \mathcal{D})$ respectively, where $W = \{W_k\}$ and $W_{(-k)}$ denotes $W$ except $W_k$.

We could describe as,

$$P(W_k|W_{(-k)}, \theta_{i_k}, \mathcal{D}) = P(W_k|W_{(-k)}, \theta_{i_k}, \mathcal{D})\mathbb{1}(\Delta_k = 0) + P(W_k|W_{(-k)}, \theta_{i_k}, \mathcal{D})\mathbb{1}(\Delta_k = 1). \tag{S3}$$

$$= P(E_k|V_{(1:N)}, \theta_{i_k}, \mathcal{D})\,(f_Y(Y_k)f_I(I_k)f_D(D_k)\mathbb{1}(\Delta_k = 0) + f_L(L_k)f_C(C_k)\mathbb{1}(\Delta_k = 1)) \tag{S4}$$

#### C. Generating $\theta$ and $W$ from their conditional posterior distributions

##### C.1. Generating $\theta$

For the prior distribution of $\theta_i$, we use a diffuse gamma distribution ($Gamma(0.001, 0.001)$) for all $i$. Then

$$p(\theta_i|W^i_{(1:n_i)}, \mathcal{D}) \propto p(\theta_i)p(W^i_{(1:n_i)}, \mathcal{D}|\theta_i),\ i \in \mathcal{A} \tag{S5}$$

when $W^i = \{W_k | \text{The age group of } k \text{ is } i\}$, $n_i$ is the population for age group $i$.

In turn, $P(W^i_1, \cdots, W^i_{n_i}, \mathcal{D}|\theta_i)$ can be expressed as

$$P(W^i_1, \cdots, W^i_{n_i}, \mathcal{D}|\theta_i) = P(E^i_1, E^i_2, \cdots, E^i_{n_i}|V_{(1:N)}, \theta_i)P(V_{(1:N)}) = \prod_{k=1}^{n_i} P(E^i_k|V_{(1:N)}, \theta_i)P(V_{(1:N)}) \tag{S6}$$

$$P(E^i_k|V_{(1:N)}, \theta_i) = P(E^i_k|I^j_{total}(t), t < E^i_k, \theta_i) = p(E^i_k|\theta_i)\prod_{t=1}^{E^i_k-1}(1 - p(t|\theta_i)) \tag{S7}$$

Since $p(E^i_k|V_{(1:N)}, \theta_i)$ is the probability of an individual $k$ to be infected at discrete time $E^i_k$ (implying the individual $k$ has not been infected before).

$$P(V_{(1:N)}) = \prod_{\Delta_k=0} 0.84 f_Y(Y_k)f_I(I_k)f_D(D_k) \prod_{\Delta_k=1} 0.16 f_L(L_k)f_C(C_k), \tag{S8}$$

where $N = \sum_i n_i$ (Total population size), $V_k = (Y_k, I_k, L_k, C_k, D_k)$ and $i_k$ is the age group of individual $k$. By applying the following approximation,

$$P(E_k|V_{(1:N)}, \theta_{i_k}) = P(E_k|I^j_{total}(t), t < E_k, \theta_{i_k}) = p(E^i_k|\theta_{i_k})\prod_{t=1}^{E^i_k-1}(1 - p(t|\theta_{i_k})) \approx p(E_k|\theta_{i_k})e^{-\sum_{t=1}^{E_k-1} p(t|\theta_{i_k})} \tag{S9}$$

we have

$$\theta_i|W, \mathcal{D} \sim Gamma\left(0.001 + |\mathcal{E}^\star_i|, 0.001 + \sum_{k \in \mathcal{E}^\star_i} \frac{\sum_{t_1}^{E_k} \phi_{ij}(t)I^j_{total}(t-1)}{n_i} + \sum_{k \in \{s: E_s > t_2 \text{ or } E_s = NA\}} \frac{\sum_{t_1}^{t_2} \phi_{ij}(t)I^j_{total}(t-1)}{n_i}\right) \tag{S10}$$

where $\mathcal{E}^\star_i$ is set of individuals in the age group $i$ exposed during the 3rd wave $[t_1, t_2]$.

##### C.2. Generating $W$

We generate $W$ by generating $W_k$ from $P(W_k|W_{(-k)}, \theta, \mathcal{D})$ iteratively, and generate $W_k$ through the Metropolis-Hasting (MH) algorithm. To sample from the posterior $P(W_k|W_{(-k)}, \theta, \mathcal{D})$ by the MH algorithm, we use the following proposal distribution:

$$Q(E_k, V_k) = Q(E_k|V_k)Q(V_k) \tag{S11}$$

$$Q(V_k) = p(V_k|\mathcal{D}) \tag{S12}$$

$$Q(E_k|V_k) = \delta(T_k - Y_k - D_k)\mathbb{1}(\Delta_k = 0) + \delta(T_k - L_k - C_k)\mathbb{1}(\Delta_k = 1) \tag{S13}$$

Putting the above together, the sampling procedure of $W_k$ is summarized in Alogorithm S1.

The acceptance ratio $\alpha$ can be obtained as follows, where $W^{(m)}_k$ and $W^{(m-1)}_k$ are denoted by $W^{(new)}_k$ and $W^{(old)}_k$, respectively. For $I_{sym}$, let $m^I_k = (T_k - D^{(new)}_k + I^{(new)}_k) \wedge (T_k - D^{(old)}_k + I^{(old)}_k)$,

**Algorithm S1.** Bayesian Inference

---

Input : $W_k^{(0)}$ for $k = 1, \cdots, N$

Sample $\theta^{(0)} = (\theta_i^{(0)})$ from prior.
**for** $m = 1 : M$(number of iteration) **do** ▷ Gibbs sampling
  **for** $k = 1 : N$ **do** ▷ MCMC
    Sample $W_k^{(m)}$ from $Q(E_k, V_k)$
    $\alpha \leftarrow \frac{P(E_k^{(m)}, E_{(-k)}^{(m-1)} | V_k^{(m)}, V_{(-k)}^{(m-1)}, \theta^{(m)}, \mathcal{D})}{P(E_k^{(m-1)}, E_{(-k)}^{(m-1)} | V_k^{(m-1)}, V_{(-k)}^{(m-1)}, \theta^{(m)}, \mathcal{D})}$ ▷ Acceptance ratio
    $W_k^{(m)} \leftarrow \begin{cases} W_k^{(m)} & \alpha \geq 1 \\ W_k^{(m)} & \text{with probability } \alpha \\ W_k^{(m-1)} & \text{else} \end{cases}$
  Sample $\{\theta_i^{(m)}\}$ for $i = 1, \cdots, A$ from $p(\theta_i | W^{(m)})$

---

$M_k^I = (T_k - D_k^{(new)} + I_k^{(new)}) \vee (T_k - D_k^{(old)} + I_k^{(old)})$. For $I_{asym}$, let $m_k^I = (T_k - C_k^{(new)}) \wedge (T_k - C_k^{(old)})$, $M_k^I = (T_k - C_k^{(new)}) \vee (T_k - C_k^{(old)})$. Here $T - D + I$ and $T - C$ are transmission onset date and $a \wedge b = \min\{a, b\}$, $a \vee b = \max\{a, b\}$.

$$\tilde{p}_i(t) = \begin{cases} p_i(t) & \text{if } t \notin [m_k^I, M_k^I] \cdots (*) \\ \frac{\phi_{ii_k}(I_{total}^{i_k}(t-1)-1)+\sum_{j \neq i_k} \phi_{ij} I_{total}^{j}(t-1)}{\phi_{ii_k}(I_{total}^{i_k}(t-1))+\sum_{j \neq i_k} \phi_{ij} I_{total}^{j}(t-1)} p_i(t) & \text{if not } (*), T_k - D_k^{(new)} + I_k^{(new)} < T_k - D_k^{(old)} + I_k^{(old)} \\ \frac{\phi_{ii_k}(I_{total}^{i_k}(t-1)+1)+\sum_{j \neq i_k} \phi_{ij} I_{total}^{j}(t-1)}{\phi_{ii_k}(I_{total}^{i_k}(t-1))+\sum_{j \neq i_k} \phi_{ij} I_{total}^{j}(t-1)} p_i(t) & \text{if not } (*), T_k - D_k^{(new)} + I_k^{(new)} > T_k - D_k^{(old)} + I_k^{(old)} \end{cases} \tag{S14}$$

when $A = \{\tilde{k} : m_k^I < E_{\tilde{k}} \leq M_k^I\}$, $B = \{\tilde{k} : E_{\tilde{k}} > M_k^I\}$

$$\alpha = \begin{cases} \frac{p_{i_k}\left(E_k^{(new)}\right)}{p_{i_k}\left(E_k^{(old)}\right)} e^{-\sum_{E_k^{(new)}}^{E_k^{(old)}-1} p_{i_k}(t)} \left(\prod_{\tilde{k} \in A} \frac{\tilde{p}_{i_{\tilde{k}}}\left(E_{\tilde{k}}\right)}{p_{i_{\tilde{k}}}\left(E_{\tilde{k}}\right)} \left(e^{\sum_{t=m_k^I}^{E_{\tilde{k}}-1} -\tilde{p}_{i_{\tilde{k}}}(t)+p_{i_{\tilde{k}}}(t)}\right)\right) \prod_{\tilde{k} \in B} \left(e^{\sum_{t=m_k^I}^{M_k^I} -\tilde{p}_{i_{\tilde{k}}}(t)+p_{i_{\tilde{k}}}(t)}\right) & E_k^{(new)} > E_k^{(old)} \\ \left(\prod_{\tilde{k} \in A} \frac{\tilde{p}_{i_{\tilde{k}}}\left(E_{\tilde{k}}\right)}{p_{i_{\tilde{k}}}\left(E_{\tilde{k}}\right)} \left(e^{\sum_{t=m_k^I}^{E_{\tilde{k}}-1} -\tilde{p}_{i_{\tilde{k}}}(t)+p_{i_{\tilde{k}}}(t)}\right)\right) \prod_{\tilde{k} \in B} \left(e^{\sum_{t=m_k^I}^{M_k^I} -\tilde{p}_{i_{\tilde{k}}}(t)+p_{i_{\tilde{k}}}(t)}\right) & E_k^{(new)} = E_k^{(old)} \\ \frac{p_{i_k}\left(E_k^{(new)}\right)}{p_{i_k}\left(E_k^{(old)}\right)} e^{\sum_{E_k^{(old)}}^{E_k^{(new)}-1} p_{i_k}(t)} \left(\prod_{\tilde{k} \in A} \frac{\tilde{p}_{i_{\tilde{k}}}\left(E_{\tilde{k}}\right)}{p_{i_{\tilde{k}}}\left(E_{\tilde{k}}\right)} \left(e^{\sum_{t=m_k^I}^{E_{\tilde{k}}-1} -\tilde{p}_{i_{\tilde{k}}}(t)+p_{i_{\tilde{k}}}(t)}\right)\right) \prod_{\tilde{k} \in B} \left(e^{\sum_{t=m_k^I}^{M_k^I} -\tilde{p}_{i_{\tilde{k}}}(t)+p_{i_{\tilde{k}}}(t)}\right) & E_k^{(new)} < E_k^{(old)} \end{cases} \tag{S15}$$

#### 2. META MODELING

We find the optimal distribution of COVID-19 vaccines for each age group to reduce the expected cases and death most, in coming 2 weeks of the study period. Let the supply of vaccine increases from 20% to 100% of total population in a stepwise manner.

Let $y_r$ be the number of the expected exposed cases (or death) in coming 2 weeks when susceptibles are decreased by vaccination proportions $r = (r_1, \ldots, r_A)^\top$. To calculate the $y_r$, we estimate $\theta_i$ by the median of the corresponding posterior distribution and impute the various necessary quantities such as exposed date, incubation period and diagnostic delay by their expected values. In addition, age-specific vaccine efficacy and fatality ratios are imputed.

The optimal vaccine proportions $r^*$ is defined as

$$r^* = (r_1^*, \cdots, r_A^*) = \arg min_r y_r \tag{S16}$$

Even though we can calculate $y_r$ for a given $r$, there is no closed equation form of $y_r$ and hence obtaining $r^*$ is challenging. We use a meta modeling approach to find $r^*$ as follows. First, we sample $r$ randomly from a Dirichlet distribution. Note that the number of vaccinated individuals at each group cannot be larger than the population size of each group and hence we accept $r$ when it satisfies this constraint. For accepted $r$s, we calculate $y_r$ to obtain pairs of $(r, y_r)$. Secondly, we fit a quadratic regression model for $(r, y_r)$, and then find a point which maximizes the fitted quadratic regression function. To maximize the quadratic function, which is not convex in general, we use the CCCP algorithm. The procedure of the proposed meta modeling is summarized as follows.

**Algorithm S2.** Meta Modeling

Input : $W_k^{(0)}$ for $k = 1, \cdots, N$

**for** $m = 1 : M$(number of iteration) **do**
    Sample $r$
    **if** $r$ satisfies the constraint **then**
        Calculate $y_r$
Using pairs of $(r, y_r)$, fit the following model

$$y = \sum_{i=1}^{A-1} \sum_{j=1}^{A-1} \alpha_{ij} r_i r_j + \sum_{i=1}^{A-1} \beta_i r_i + \epsilon \tag{S17}$$

▷ Since the sum of $r$ is 1, we remove a $r_i$ when fitting

By using CCCP algorithm, we optimize the following QP problem.

$$r^* = \arg min_r \sum_{i=1}^{A-1} \sum_{j=1}^{A-1} \hat{\alpha}_{ij} r_i r_j + \sum_{i=1}^{A-1} \hat{\beta}_i r_i \tag{S18}$$

when, $1 - \frac{n_i}{N_{vac}} \leq \sum_{i=1}^{A-1} r_i \leq 1, 0 \leq r_i \leq \frac{n_i}{N_{vac}}$.
