## Supplementary figures and images for "COVID-19 Vaccine Prioritisation in Japan and South Korea"

### Supplementary Figure

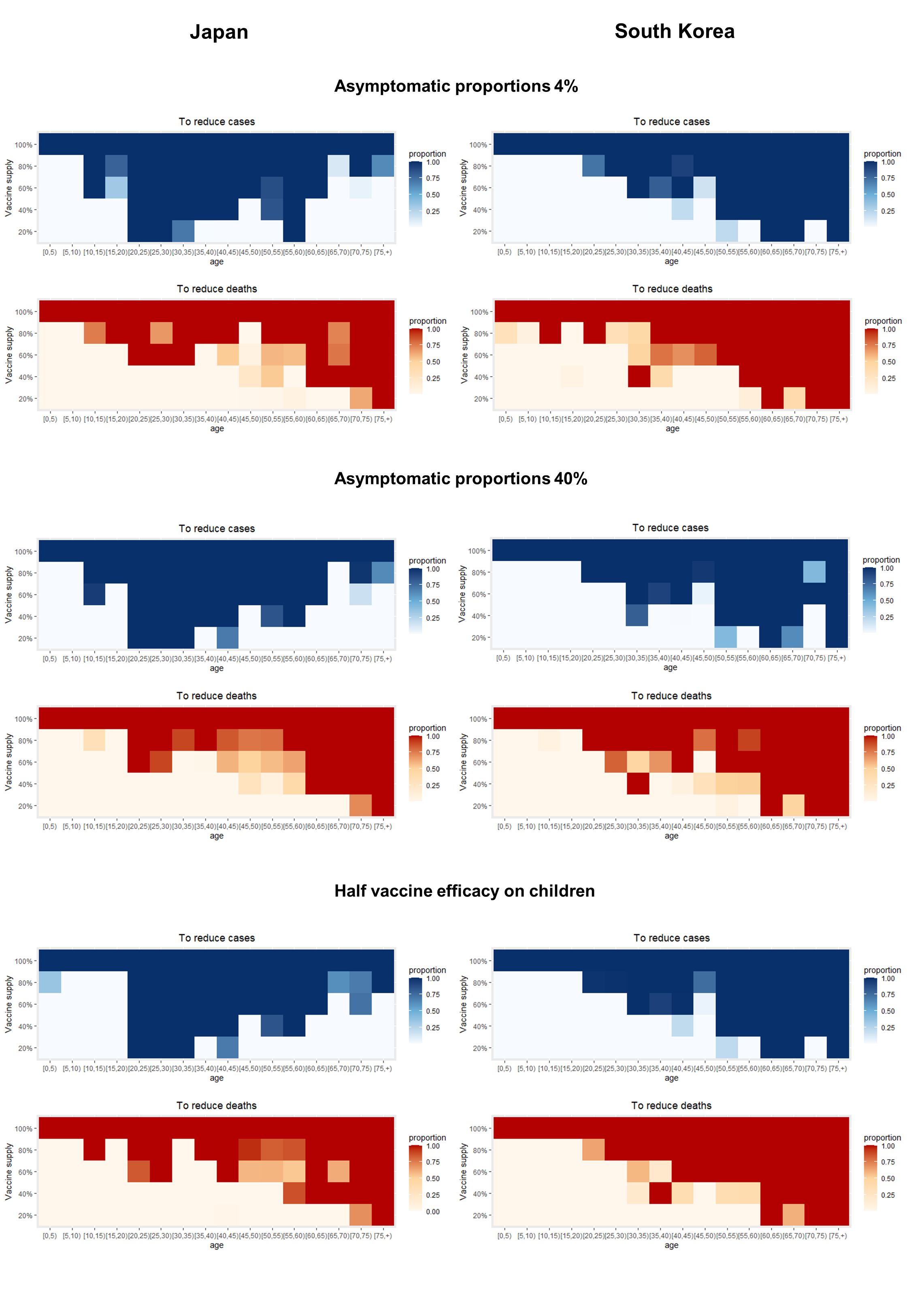
